# Cell-based therapies for COVID-19: A living systematic review

**DOI:** 10.1101/2020.04.24.20078667

**Authors:** Gabriel Rada, Javiera Corbalán, Patricio Rojas

**Author notes:** **Corresponding author:** Gabriel Rada, Email address, Postal address: Holanda 895, Providencia, Santiago, Chile. **COVID-19 L·OVE Working Group**.

## Abstract

**Objective:** This living systematic review aims to provide a timely, rigorous and continuously updated summary of the available evidence on the role of cell-based therapies in the treatment of patients with COVID-19.

**Data sources:** We conducted searches in PubMed/Medline, Embase, Cochrane Central Register of Controlled Trials (CENTRAL), grey literature and in a centralised repository in L-OVE (Living OVerview of Evidence). L-OVE is a platform that maps PICO questions to evidence from Epistemonikos database. In response to the COVID-19 emergency, L-OVE was adapted to expand the range of evidence it covers and customised to group all COVID-19 evidence in one place. All the searches covered the period until April 23, 2020 (one day before submission).

**Eligibility criteria for selecting studies and methods:** We adapted an already published common protocol for multiple parallel systematic reviews to the specificities of this question. We searched for randomised trials evaluating the effect of cell-based therapies versus placebo or no treatment in patients with COVID-19. Anticipating the lack of randomised trials directly addressing this question, we also searched for trials evaluating other coronavirus infections, such as MERS-CoV and SARS-CoV, and non-randomised studies in COVID-19. Two reviewers independently screened each study for eligibility.

A living, web-based version of this review will be openly available during the COVID-19 pandemic. We will resubmit it every time the conclusions change or whenever there are substantial updates.

**Results:** We screened 1,043 records but no study was considered eligible.

We identified 61 ongoing studies, including 39 randomised trials evaluating different types of cell-based therapies in COVID-19.

**Conclusions:** We did not find any studies that met our inclusion criteria and hence there is no evidence to support or refute the use of cell-based therapies in the treatment of patients with COVID-19. A substantial number of ongoing studies should provide valuable evidence to inform researchers and decision makers in the near future.

**PROSPERO Registration number:** CRD42020179711

Box 1

Linked resources in this Living Systematic Review Common protocol
Common protocol for the systematic reviews and overviews of systematic reviews being conducted by the COVID-19 L·OVE Working Group.
[Available here](http://dx.doi.org/10.5867/medwave.2020.03.7867)

Living review
Web version of this systematic review, presented in a ‘living systematic review format’. This means it is continuously updated as soon as new evidence emerges.
[Available here](https://app.iloveevidence.com/living-reviews/cell-based-therapies-for-covid-1)

Living OVerview of Evidence - L·OVE
An open platform that uses artificial intelligence and a broad network of contributors to identify all of the evidence relevant to this and other healthcare questions, including those related to COVID-19.
[Available here](https://app.iloveevidence.com/loves/5e6fdb9669c00e4ac072701d?question_domain=5b1dcd8ae611de7ae84e8f14□population=5e6fdab269c00e4ac072701c□intervention=5e973bce3552583c298cca05)

## INTRODUCTON

COVID-19 is an infection caused by the SARS-CoV-2 coronavirus [1]. It was first identified in Wuhan, China, on December 31, 2019 [2]; By April 23, 2020, the number of confirmed COVID-19 cases had reached 2,761,121, with 193,671 confirmed deaths [3]. On March 11, 2020, WHO characterised the COVID-19 outbreak as a pandemic [1].

While the majority of cases result in mild symptoms, some might progress to pneumonia, acute respiratory distress syndrome, and death [4],[5],[6]. The case fatality rate reported across countries, settings and age groups is highly variable, but it ranges from about 0.5% to 10% [7]. In some centres it has been reported to be higher than 10% in hospitalised patients [8].

Cell-based therapy is a treatment in which viable nucleated cells are injected, grafted or implanted into a patient in order to obtain a medicinal effect. Mesenchymal stem cells are the main type of cell-based therapy proposed in the context of COVID-19, but many others are being actively researched [9]. Mesenchymal stem cells exhibit a capacity for homing to sites of injury and inflammation where they exert antiinflammatory and immunomodulatory effects [10]. They can affect the status of T cells and skew them towards a regulatory phenotype, and they also interact with B cells, inhibiting B cell response [11].

Preclinical studies in COVID-19 patients suggest mesenchymal stromal cells would be able to reduce inflammation [12]. Several findings support this hypothesis, such as the expression of antiinflammatory and trophic factors, and the decrease of C-reactive protein, tumor necrosis factor-alpha, and cytokine-secreting immune cells [13].

In consequence, it is expected that these cells could attenuate the overactivation of the immune system and support repair by modulating the lung microenvironment after SARS-CoV-2 infection. Some small studies in humans and newspaper reports have sparked global interest in this treatment, but high-quality evidence is still lacking [14].

Using innovative and agile processes, taking advantage of technological tools, and resorting to the collective effort of several research groups, this living systematic review aims to provide a timely, rigorous and continuously updated summary of the available evidence on the role of cell-based therapies in the treatment of patients with COVID-19.

## METHODS

This manuscript complies with the ‘Preferred Reporting Items for Systematic reviews and Meta-Analyses’ (PRISMA) guidelines for reporting systematic reviews and meta-analyses [15] (see **Appendix 1- PRISMA Checklist)**.

A protocol stating the shared objectives and methodology of multiple evidence syntheses (systematic reviews and overviews of systematic reviews) to be conducted in parallel for different questions relevant to COVID-19 was published elsewhere [16]. The review was registered in PROSPERO with the number CRD42020179711 and a detailed protocol was uploaded to a preprint server [17]

### Search strategies

#### Electronic searches

Our literature search was devised by the team maintaining the L·OVE platform (https://app.iloveevidence.com), using the following approach:

1. Identification of terms relevant to the population and intervention components of the search strategy, using Word2vec technology [18] to the corpus of documents available in Epistemonikos Database.
2. Discussion of terms with content and methods experts to identify relevant, irrelevant and missing terms.
3. Creation of a sensitive boolean strategy encompassing all the relevant terms
4. Iterative analysis of articles missed by the boolean strategy, and refinement of the strategy accordingly.

Our main search source was Epistemonikos database (https://www.epistemonikos.org), a comprehensive database of systematic reviews and other types of evidence [19] that we have supplemented it with information coming from 35 sources relevant to COVID-19. The list of sources that have been added to Epistemonikos Database is continuously expanded. This list of sources regularly screened by Epistemonikos for COVID-19 is [updated regularly in our website](https://app.iloveevidence.com/covid19/methods) [20].

We conducted additional searches using highly sensitive searches in PubMed/MEDLINE, the Cochrane Central Register of Controlled Trials (CENTRAL) and Embase.

The searches in Epistemonikos are continuously updated [20] but were last checked the day of submission of this article (April 24, 2020). The additional searches were updated on April 23, 2020, and covered the period from the inception date of each database. No study design, publication status or language restriction were applied to the searches in Epistemonikos or the additional electronic searches.

The following strategy was used to search in Epistemonikos Database. We adapted it to the syntax of other databases (see **Appendix 2- Search strategies)**.

((coronavir* OR coronovirus* OR “corona virus” OR “virus corona” OR “corona virus” OR “virus corona” OR hcov* OR “covid-19” OR covidl9* OR “covid 19” OR “2019-nCoV” OR cvl9* OR “cv-19” OR “cv 19” OR “n-cov” OR ncov* OR “sars-cov-2” OR “sars-cov2” OR “SARS-Coronavirus-2” OR “SARS-Coronavirus2” OR (wuhan* AND (virus OR viruses OR viral)) OR (covid* AND (virus OR viruses OR viral)) OR “sars-cov” OR “sars cov” OR “sars-coronavirus” OR “severe acute respiratory syndrome” OR “mers-cov” OR “mers cov” OR “middle east respiratory syndrome” OR “middle-east respiratory syndrome” OR “covid-19-related” OR “SARS-CoV-2-related” OR “SARS-CoV2-related” OR “2019-nCoV-related” OR “cv-19-related” OR “n-cov-related”)) AND (“cell therapy” OR “cell therapies” OR “cell-therapy” OR “cell-therapies” OR “mesenchymal cell” OR “mesenchymal cells” OR MSC OR MSCs OR HMSC* OR stemstromal* OR stromalstem* OR nestcell* OR ((mesenchymal* OR “tissue-derived” OR “derived-mesenchymal”) AND (stromal* OR stem OR multipotent* OR progenitor*)) OR (medicinal* AND signalling* AND (cell OR cells)) OR (stromal* AND (stem OR multipotent*)) OR (“tissue-derived” AND mesenchymal*))

#### Other sources

In order to identify articles that might have been missed in the electronic searches, we proceed, if necessary, as follows:

1. Screened the reference lists of other systematic reviews.
2. Scanned the reference lists of selected guidelines, narrative reviews and other documents.
3. Reviewed websites specialised in COVID-19.
4. Email the contact authors of all the included studies to ask for additional publications or data on their studies, and for other studies in the topic.
5. Conduct cross-citation searches in Google Scholar and Microsoft Academic, using each included study as the index reference.
6. Review the reference list of each included study.

### Eligibility criteria

#### Types of studies

This living review preferentially includes randomised trials. Non-randomised studies are included if there is no direct evidence from randomised trials or the certainty of evidence for the critical outcomes resulting from the randomised trials is graded as low- or very low, and the certainty provided by the non-randomised evidence grades higher than the one provided by the randomised evidence [21].

We exclude studies evaluating the effects on animal models or in vitro conditions.

#### Types of participants

We include trials assessing participants with COVID-19, as defined by the authors of the trials. Whenever we find substantial clinical heterogeneity on how the condition was defined, we explore it using a sensitivity analysis.

If we do not find direct evidence from randomised trials, or if the evidence from randomised trials provided low- or very low-certainty evidence for critical outcomes, we consider eligible randomised trials evaluating cell-based therapy in other coronavirus infections, such as MERS-CoV or SARS-CoV infections [21].

#### Type of interventions

The interventions of interest are cell-based therapies obtained from any tissue, including mesenchymal stromal cells, hematopoietic stem cells, and any other therapy in which viable nucleated cells are injected, grafted or implanted into a patient in order to obtain a medicinal effect.

We do not restrict our criteria to any dosage, duration, timing or route of administration. The comparison of interest is placebo/sham (cell-based therapy plus optimal treatment versus placebo plus optimal treatment) or no treatment (cell-based therapy plus optimal treatment versus optimal treatment).

Trials assessing cell-based therapy plus other interventions are eligible if the cointerventions are identical in both the intervention and the comparison groups. Trials evaluating cell-based therapy in combination with other active interventions versus placebo or no treatment are also eligible.

#### Type of outcomes

We do not use the outcomes as an inclusion criteria during the selection process. Any article meeting all the criteria except for the outcome criterion is preliminarily included and assessed in full text.

We used the core outcome set COS-COVID [22], the existing guidelines and reviews and the judgement of the authors of this review as an input for selecting the primary and secondary outcomes, as well as to decide upon inclusion. The review team regularly revise this list of outcomes, in order to incorporate ongoing efforts to define Core Outcomes Sets (e.g. COVID-19 Core Outcomes [23].

##### Primary outcome

- All-cause mortality

##### Secondary outcomes

- Mechanical ventilation
- Extracorporeal membrane oxygenation
- Length of hospital stay
- Respiratory failure
- Serious adverse events
- Time to SARS-CoV-2 RT-PCR negativity

##### Other outcomes

- Acute respiratory distress syndrome
- Total adverse events

If we include at least one study, primary and secondary outcomes are presented in the GRADE ‘Summary of Findings’ tables, and a table with all the outcomes is presented as an appendix [24].

### Selection of studies

The results of the literature search in Epistemonikos database are automatically incorporated into the [L·OVE platform](https://test.iloveevidence.com/loves/5e6fdb9669c00e4ac072701d) (automated retrieval). There they are de-duplicated by an algorithm that compares unique identifiers (database ID, DOI, trial registry ID), and citation details (i.e. author names, journal, year of publication, volume, number, pages, article title, and article abstract).

The additional searches are uploaded to the screening software Collaboratron™[25].

In both L·OVE platform and Collaboratron™, two researchers independently screen the titles and abstracts yielded by the search against the inclusion criteria. We obtain the full reports for all titles that appear to meet the inclusion criteria or require further analysis, and then decide about their inclusion.

We record the reasons for excluding trials in any stage of the search and outline the study selection process in a PRISMA flow diagram which we adapted for the purpose of this project.

### Extraction and management of data

Using standardised forms, two reviewers independently extract data from each included and ongoing study. We collect the following information: study design, setting, participant characteristics (including disease severity and age) and study eligibility criteria; details about the administered intervention and comparison, including source of cells, dose, duration and timing (i.e. time after diagnosis); the outcomes assessed and the time they were measured; the source of funding of the study and the conflicts of interest disclosed by the investigators; the risk of bias assessment for each individual study.

We resolve disagreements by discussion, and one arbiter adjudicates unresolved disagreements.

### Risk of bias assessment

The risk of bias for each randomised trial is assessed by using the ‘risk of bias’ tool (RoB 2.0: a revised tool to assess risk of bias in randomised trials) [26]. We consider the effect of assignment to the intervention for this review. Two reviewers independently assess five domains of bias for each outcome result of all reported outcomes and time points. These five domains are: bias due to (1) the randomisation process, (2) deviations from intended interventions (effects of assignment to interventions at baseline), (3) missing outcome data, (4) measurement of the outcome, and (5) selection of reported results. Answers to signalling questions and collectively supporting information leads to a domain-level judgement in the form of ‘Low risk of bias’, ‘Some concerns’, or ‘High risk of bias’. These domain-level judgements inform an overall ‘risk of bias’ judgement for each result. Discrepancies between review authors are resolved by discussion to reach consensus. If necessary, a third review author is consulted to achieve a decision.

We assess the risk of bias of other study designs with the ROBINS-I tool (ROBINS-I: Risk Of Bias In Non-randomised Studies of Interventions) [27]. We address the following domains: bias due to confounding, bias in selection of participants into the study, bias in classification of interventions, bias due to deviations from intended interventions (effect of assignment to intervention), bias due to missing data, bias in measurement of outcomes and bias in the selection of the reported result. We judge each domain as low risk, moderate risk, serious risk, critical risk, or no information, and evaluate individual bias items as described in ROBINS-I guidance. We do not consider time-varying confounding, as these confounders are not relevant in this setting [28].

We consider the following factors as baseline potential confounders:

- Age
- Comorbidities (e.g. cardiovascular disease, renal disease, eye disease, liver disease)
- Co-interventions
- Severity, as defined by the authors (i.e respiratory failure vs respiratory distress syndrome vs ICU requirement).

### Measures of treatment effect

For dichotomous outcomes, we express the estimate of treatment effect of an intervention as risk ratios (RR) or odds ratios (OR) along with 95% confidence intervals (CI).

For continuous outcomes, we use the mean difference and standard deviation (SD) to summarise the data using a 95 % CI. Whenever continuous outcomes are measured using different scales, the treatment effect is expressed as a standardised mean difference (SMD) with 95% CI. When possible, we multiply the SMD by a standard deviation that is representative from the pooled studies, for example, the SD from a well-known scale used by several of the studies included in the analysis on which the result is based. In cases where the minimally important difference (MID) is known, we present continuous outcomes as MID units or inform the results as the difference in the proportion of patients achieving a minimal important effect between intervention and control [28]. Then, these results are displayed on the ‘Summary of Findings Table’ as mean difference [28].

### Strategy for data synthesis

If we include more than one trial, we conduct meta-analysis for studies clinically homogeneous using RevMan 5 [29], using the inverse variance method with random effects model. For any outcomes where data are insufficient to calculate an effect estimate, a narrative synthesis is presented.

### Subgroup and sensitivity analysis

We perform subgroup analysis according to the definition of severe COVID-19 infection (i.e respiratory failure vs respiratory distress syndrome vs ICU requirement). In case we identify significant differences between subgroups (test for interaction <0.05) we report the results of individual subgroups separately.

We perform sensitivity analysis excluding high risk of bias studies, and if non-randomised studies are used, excluding studies that do not report adjusted estimates. In cases where the primary analysis effect estimates and the sensitivity analysis effect estimates significantly differ, we either present the low risk of bias – adjusted sensitivity analysis estimates – or present the primary analysis estimates but downgrading the certainty of the evidence because of risk of bias.

### Assessment of certainty of evidence

The certainty of the evidence for all outcomes is judged using the Grading of Recommendations Assessment, Development and Evaluation working group methodology (GRADE Working Group) [30], across the domains of risk of bias, consistency, directness, precision and reporting bias. Certainty is adjudicated as high, moderate, low or very low. For the main comparisons and outcomes, we prepare Summary of Findings (SoF) tables [28],[31] and also interactive Summary of Findings tables (http://isof.epistemonikos.org/). A SoF table with all the comparisons and outcomes is presented as an appendix.

### Living evidence synthesis

An artificial intelligence algorithm deployed in the [Coronavirus/COVID-19 topic of the L·OVE platform](https://app.iloveevidence.com/loves/5e6fdb9669c00e4ac072701d) provides instant notification of articles with a high likelihood of being eligible. The authors review them, decide upon inclusion, and update the [living web version](https://app.iloveevidence.com/living-reviews/5e7e748c53e53e78053563fd) of the review accordingly. We expect to resubmit to a journal any time there is a change in the direction of the effect on the critical outcomes or a substantial modification to the certainty of the evidence.

This review is part of a larger project set up to produce multiple parallel systematic reviews relevant to COVID-19 [16].

## RESULTS

### Results of the search

We used a repository that includes searches in 34 trial registries, pre-print servers and websites specialised in COVID-19. We also conducted additional searches in 3 electronic databases and scanned the references of multiple guidelines, reviews and other documents.

The search in the L·OVE platform retrieved 272 records, and the additional searches retrieved 771 records (total records screened= 1 043). We considered 78 as potentially eligible and retrieved and evaluated their full texts. However, none of the studies were eligible for inclusion. The reasons for exclusion are described in the **Appendix 3 - List of relevant studies**

The study selection process is summarised in Figure 1 - PRISMA Flowchart.

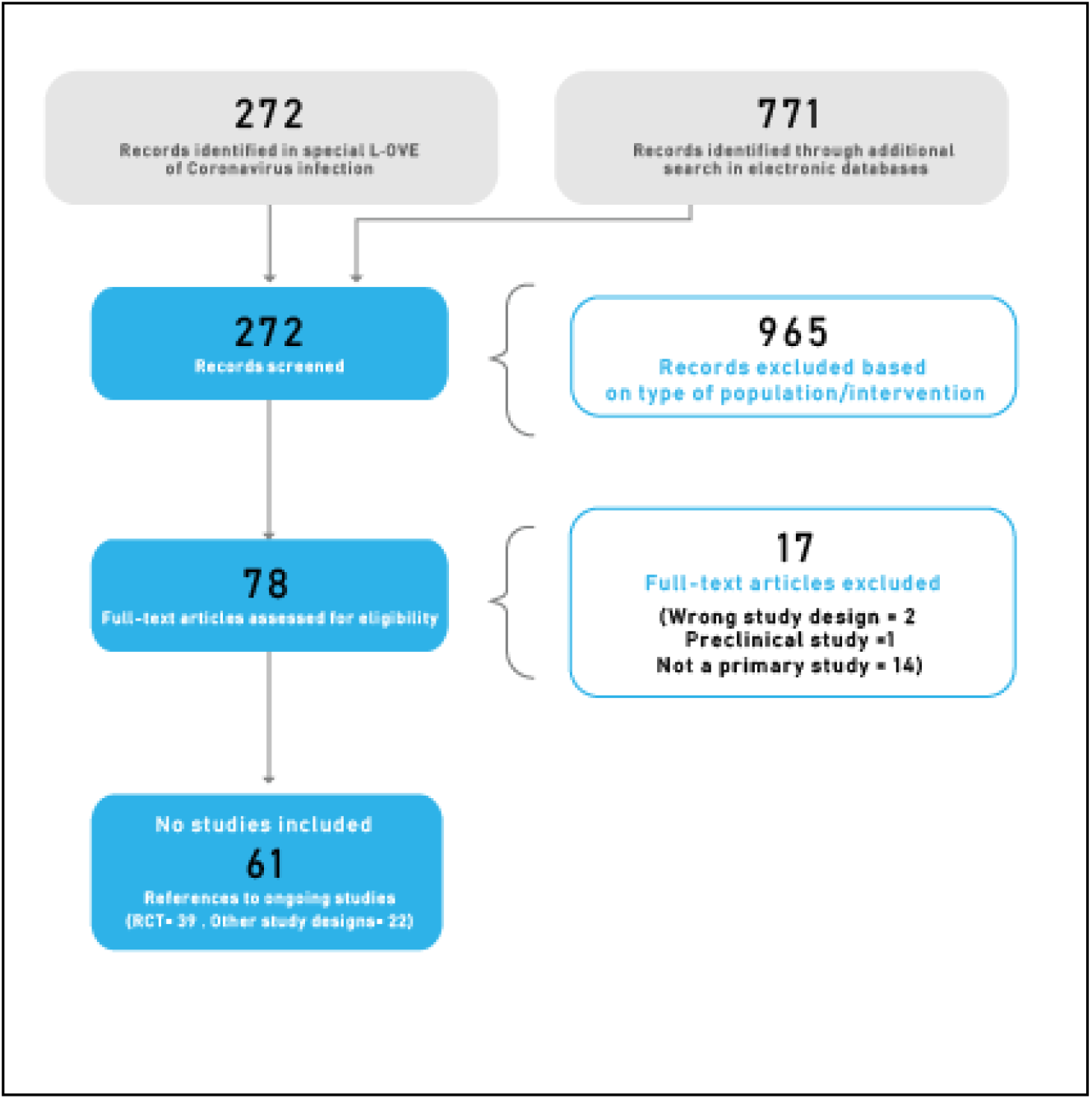

### Description of the studies

No study was considered eligible.

### Ongoing studies

We identified 61 ongoing studies (39 randomised trials and 22 non-randomised studies). See **Appendix 3 - List of relevant studies**.

## DISCUSSION

We performed a comprehensive search of the literature and did not find any randomised trials evaluating the effect of cell-based therapies in patients with COVID-19. Anticipating this lack of randomised trials, we also searched for non-randomised, comparative studies in COVID-19 and for randomised trials evaluating other coronavirus infections, such as MERS-CoV and SARS-CoV. These additional searches provided no relevant studies either. In sum, we did not find any studies fulfilling the minimum requirements to inform decisions.

Systematic reviews are the gold standard to collect and summarize the available evidence regarding a scientific question. However, the traditional model for conducting reviews has several limitations, including high demand for time and resources [34] and a rapid obsolescence [35]. Amidst the COVID-19 crisis, researchers should make their best effort to answer the urgent needs of health decision makers yet without giving up scientific accuracy. Information is being produced at a vertiginous speed [36], so alternative models are needed.

One potential solution to these shortfalls are rapid reviews, reviews that omit some of the steps of a traditional systematic review in order to move faster. Unfortunately, in these reviews rapidity comes at the cost of quality [37]. Furthermore, they do not solve the issue of obsolescence. Living systematic reviews do address that issue [38]. They are continually updated by incorporating relevant new evidence as it becomes available, at a substantial effort. So, an approach combining these two models might prove more successful in providing the scientific community and other interested parties with evidence that is actionable, rapidly and efficiently produced, up to date, and of the highest quality [39].

This review is part of a larger project set up to put such an approach into practice. The project aims to produce multiple parallel living systematic reviews relevant to COVID-19 following the higher standards of quality in evidence synthesis production [19]. We believe our methods are well suited to handle the abundance of evidence that is to come, including evidence on the role of cell-based therapies in the treatment of patients with COVID-19. We have identified multiple ongoing studies addressing this question, including 39 randomised trials, which will provide valuable evidence to inform researchers and decision makers in the near future.

We found two systematic reviews, with a broad scope, addressing the question of this article [40],[41]. Their conclusions are similar in terms of the lack of evidence available. So, the main limitation of the existing systematic reviews, including ours, results from the absence of any evidence to inform decisions. Nonetheless, decisions should be made even if the evidence is considered insufficient [42. While the evidence about the potential benefit emerges, decision makers will have to ponder other relevant aspects [43].

We think the main factors that decision makers must put in the balance are the high costs of this treatment. On the other hand, researchers in this field should deepen our understanding about how cell-therapies work and which specific therapy is the better option. We hope that the high number of studies that are expected to be completed in the next months will shed some light on these issues.

During the COVID-19 pandemic we will maintain a living, web-based, openly available version of this review, and we will resubmit the review every time the conclusions change or whenever there are substantial updates. Our systematic review aims to provide a high-quality, up-to-date synthesis of the evidence that is useful for clinicians and other decision makers.

## Data Availability

All data related to the project will be available. Epistemonikos Foundation will grant access to data.

## NOTES

### Differences between protocol and review

Our original protocol intended to include studies evaluating mesenchymal stem cells, the main type of cell-based therapy proposed in the context of COVID-19. In consultation with experts in the field, we expanded our criteria to include other types of cell-based therapies as well.

## Acknowledgements

The members of the COVID-19 L·OVE Working Group and [Epistemonikos Foundation](https://www.epistemonikos.cl/) have made possible to build the systems and compile the information needed by this project. Epistemonikos is a collaborative effort, based on the ongoing volunteer work of over a thousand contributors since 2012.

## Roles and contributions

GR conceived the common protocol for all the reviews being conducted by the COVID-19 L·OVE Working Group. GR drafted the manuscript, and all other authors contributed to it. The corresponding author is the guarantor and declares that all authors meet authorship criteria and that no other authors meeting the criteria have been omitted.

The COVID-19 LOVE Working Group was created by Epistemonikos and a number of expert teams in order to provide decision makers with the best evidence related to COVID-19. Up-to-date information about the group and its member organisations is available here: epistemonikos.cl/working-group

## PROSPERO registration number

CRD42020179711

## Ethics

As researchers will not access information that could lead to the identification of an individual participant, obtaining ethical approval was waived.

